# Frostbite Immersion Rewarming Methods: Sink & Faucet vs Bucket vs Immersion Circulator

**DOI:** 10.1101/2024.11.04.24316457

**Authors:** Alberta Frostbite Care Collaborative, Matthew J. Douma, Jaskirat D. Tiwana

## Abstract

Frostbite can lead to cellular damage, vascular injury, and an altered functional status. Current rapid rewarming methods are unreliable at maintaining water temperature between the gold standard range of 37°C and 39°C. However, immersion circulators can precisely maintain this temperature range throughout rewarming, potentially leading to better patient outcomes following frostbite injury. In-vitro rewarming trials were conducted using frozen wild boar (*Sus scrofa*) legs to evaluate 3 rewarming methods: sink and faucet, bucket, and immersion circulator. Porcine leg temperature and water temperature were measured every minute over the 30 minute duration of each trial. The immersion circulator method proved to be the most efficient at reaching the target tissue temperature (9 minutes) while maintaining the least variability in water temperature (0% of time spent below 37°C). Our study concludes that the immersion circulator method is superior to other methods as it achieves faster and more consistent rewarming. This method has the potential to enhance frostbite treatment protocols, particularly in clinical and field settings where consistent rewarming is difficult to achieve.

---

Frostbite is a localized cold injury caused by prolonged exposure to freezing temperatures, resulting in ice crystal formation within cells and tissues, leading to cellular damage, vascular injury, and potentially necrosis (Imray et al., 2009). Damage occurs in phases, beginning with vasoconstriction and tissue cooling, followed by cellular ice formation and thawing, which triggers inflammation and vascular collapse (Imray et al., 2009). The extent of frostbite injury depends on several factors, including the duration of cold exposure, the rapidity of tissue freezing, and the rewarming method used (McIntosh et al., 2011). Rapid rewarming in warm water between 37°C and 39°C remains the gold standard for frostbite treatment, as it mitigates further tissue damage by promoting vasodilation, improving microvascular perfusion, and restoring oxygen to the tissues (Handford et al., 2014; Heggers et al., 1987).

Achieving consistent, precise rewarming is critical because improper techniques can lead to refreezing, thermal injury, or exacerbation of ischemic injury. Particularly in resource-limited settings or field environments, maintaining the ideal temperature for rewarming is challenging (McIntosh et al., 2011). Emerging technologies, such as immersion circulator devices, may offer a promising solution by allowing for precise control of water temperature, potentially minimizing tissue damage and improving outcomes. This study compares three rewarming methods—faucet/sink, bucket, and immersion circulator—to evaluate their effectiveness, consistency, and speed in rewarming frostbitten tissue. We hypothesize that the immersion circulator method, due to its precise temperature control, will provide the fastest and most consistent rewarming with the least variability, thereby reducing the risk of further tissue injury.

## Clinical Context and Rationale

The primary goal of frostbite treatment is to restore blood flow and oxygenation to the affected tissues as rapidly as possible to prevent necrosis and limit complications such as infection, compartment syndrome, and amputation (McIntosh et al., 2011). Warm water immersion, between 37°C and 39°C, is the most effective rewarming strategy because it optimizes vasodilation, enhances microvascular circulation, and prevents the further formation of ice crystals in the tissues (Heggers et al., 1987). Controlled, rapid rewarming has been associated with reduced amputation rates and improved long-term functional outcomes in frostbite patients (Handford et al., 2014).

However, maintaining this precise temperature range is difficult, especially in prehospital, field, or emergency department settings, where environmental conditions and resource constraints may limit the ability to perform optimal rewarming (Imray et al., 2009). The use of immersion circulator technology, which allows for precise and stable temperature control, may overcome these challenges. Originally developed for culinary purposes, immersion circulator devices are capable of maintaining water temperature to within fractions of a degree, offering a potentially valuable tool in the clinical management of frostbite. By providing stable rewarming, immersion circulator devices may improve tissue salvage, reduce complications, and offer a more consistent rewarming method than traditional approaches.

### Purpose of the Simulation Study

The primary objective of this study is to evaluate the performance of different rewarming techniques in achieving the target tissue temperature of 38°C in a controlled setting, using pig legs as a model for frostbitten tissue. The goal is to assess which method provides the most effective and consistent rewarming, thereby reducing the risk of further tissue damage.

## Methods

### Specimen Preparation

Porcine legs (*Sus scrofa*) were obtained from a local supplier and immediately instrumented with temperature probes for monitoring. The legs were then placed in a laboratory freezer and cooled to -8°C to simulate frostbite conditions.

### Study Procedure

Prior to the start of each rewarming trial, the legs were removed from the freezer and placed in a 38°C rewarming environment. Each trial commenced immediately upon placing the frozen leg into the water, and temperature measurements were monitored continuously and recorded every minute throughout the rewarming process to monitor progress. Two sources of temperature data were collected during each trial: in-tissue temperatures and rewarming water temperatures. The study protocol required that the rewarming water be maintained at 38°C.

### Rewarming Methods

Three different rewarming methods were prepared and evaluated.

#### Method #1: Continuously running sink

The first method involved running warm water from a faucet into a partially filled sink, where the tap water was set to 38°C. The water ran continuously and the sink drain was left partially open, allowing for a consistent water level to be maintained during the 30 minute rewarming trial.

#### Method #2: Multiple bucket method

The second method utilized a 15-liter stainless steel basin that was initially filled with 5 liters of warm water at 38°C. As the water temperature dropped during the rewarming process, cooler water was removed from the basin using a graduated cylinder and additional warm water was added to maintain the target rewarming temperature.

#### Method #3: Immersion circulator

The final method employed an immersion circulator, which was set to maintain a water temperature of 38°C. The device was placed in a water bath, allowing it to automatically regulate the water temperature without the need for manual intervention.

### Equipment

For monitoring tissue temperature, the ThermoPro TP25 Wireless Meat Thermometer was used. The water temperature was measured using the Inkbird IBS-TH2 Plus Bluetooth Temperature Probe. The immersion circulator used was the PolyScience MX-CA11B, chosen because it is a laboratory and industrial quality precision circulator approved for use in high-sensitivity applications. For the bucket trial, a medical grade stainless steel basin with measurements of length 37.5cm, width 20cm, and depth 20cm was used.

### Data Recording

All temperature data was manually recorded in Microsoft Excel for each trial. The data included the initial water temperature, the tissue temperature at the start of rewarming, and recordings taken every minute for the duration of the trial.

### Statistical Analysis

Statistical analysis was performed using R statistical software. One-way ANOVA was used to compare mean rewarming temperatures between the three methods, followed by Tukey’s Honest Significant Difference (HSD) test for pairwise comparisons. Descriptive statistics, including means, standard deviations, and ranges, were calculated for each rewarming method. The coefficient of variation (CV) was also calculated to assess the variability in temperature control.

### Visualization

Visualizations, including line graphs to depict temperature changes over time for each method, were created using Python’s Matplotlib library. These graphs visually demonstrated the differences in temperature stability and rewarming efficiency across the three methods. The area under the curve (AUC) was calculated to assess cumulative rewarming efficiency.

### Ethical Considerations

Ethics approval was not required for this study, as it involved non-human, non-living animal tissue, in accordance with the institutional guidelines for research involving animal byproducts.

### Team Composition

The trial team consisted of three members:

1. A supervisor who oversaw the experiment and ensured adherence to the protocol.
2. A technician emergency registered nurse with >10 years experience, responsible for conducting the rewarming trial.
3. A recorder who logged temperature measurements and managed data recording in Excel.

## Results

### Descriptive Statistics

The three rewarming methods—faucet/sink, bucket, and immersion circulator—were assessed based on their effectiveness in rewarming pig leg tissue to a target temperature of 38°C over a 30-minute period. The results, summarized in Table 1, provide descriptive statistics for water temperature across all three methods.

**Table 1.** Descriptive Statistics for Water Temperature by Rewarming Method.

| Method | Mean Temp (°C) | SD (°C) | Min Temp (°C) | Max Temp (°C) |
| --- | --- | --- | --- | --- |
| Faucet/sink | 36.30 | 1.22 | 34.16 | 38.84 |
| Bucket | 36.31 | 1.84 | 33.12 | 39.45 |
| Immersion Circulator | 38.03 | 0.11 | 37.84 | 38.18 |

**Table 2.** Water Temperature Below Therapeutic Rewarming Window by Rewarming Method.

| Method | Time below 37°C (minutes) | Percentage of time below 37°C (%) |
| --- | --- | --- |
| Faucet/sink | 21 | 70.0 |
| Bucket | 23 | 76.7 |
| Immersion Circulator | 0 | 0.0 |

*Note.* Highlights of Figure 1.

The faucet/sink method demonstrated a mean water temperature of 36.30°C (SD = 1.22°C), with a wide range of temperatures fluctuating between 34.16°C and 38.84°C, indicating significant variability in temperature control. The bucket method showed even greater variability, with a mean water temperature of 36.31°C (SD = 1.84°C) and a temperature range of 33.12°C to 39.45°C. The need for frequent manual intervention to maintain the target temperature contributed to this high level of variability.

In contrast, the immersion circulator method maintained the most consistent temperature, with a mean of 38.03°C (SD = 0.11°C) and a range of 37.84°C to 38.18°C. This method exhibited significantly less variability compared to the other two, providing superior temperature stability throughout the rewarming process.

### Time to Target Temperature

The time required to reach the target tissue temperature of 38°C varied across the methods (Figure 1). The immersion circulator method achieved the target temperature in approximately 9 minutes, demonstrating the fastest rewarming performance. The bucket method reached 38°C in approximately 22 minutes, while the faucet/sink method was the slowest, taking approximately 29 minutes to rewarm the tissue.

**Figure 1.**
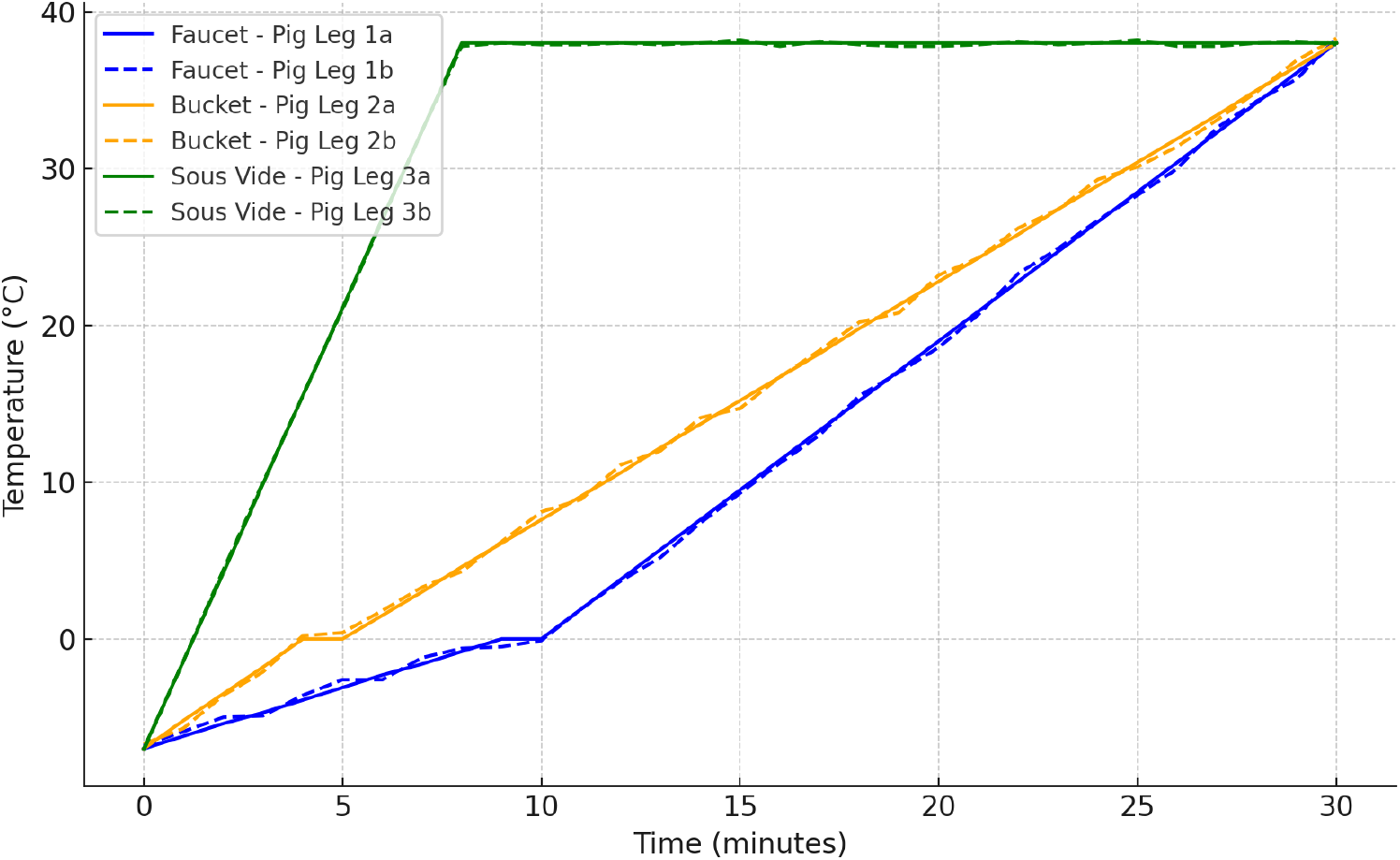
Time to reach target tissue temperature for faucet/sink, bucket, and immersion circulator (sous vide) methods.

### Rewarming Water Variability

### Statistical Comparisons

#### One-Way ANOVA

A one-way analysis of variance (ANOVA) was conducted to compare the mean rewarming temperatures across the three methods. The results revealed a significant difference in mean temperatures between the methods: F(2, 180) = 290044.81, p < 0.001. This indicates that the rewarming performance differed significantly among the faucet/sink, bucket, and immersion circulator methods.

#### Tukey’s HSD Post-Hoc Test

To further explore the differences between methods, Tukey’s Honest Significant Difference (HSD) post-hoc test was applied. The results showed significant differences in mean temperatures between all pairs of methods, all p values < 0.05. These results confirm that the immersion circulator method is significantly faster and more consistent in rewarming compared to both the faucet/sink and bucket methods.

#### Variability in Temperature Control

As shown in Figure 2, the faucet/sink and bucket methods exhibited substantial temperature fluctuations. The faucet/sink method had water temperatures fluctuating between 34°C and 39°C, including water temperature fluctuations coming from the tap, while the bucket method showed even greater variability, with frequent drops in temperature that required manual intervention, nearly continuously, to rewarm the water. In contrast, the immersion circulator method maintained precise temperature control with minimal deviation from the target temperature (38°C ± 0.18°C).

**Figure 2.**
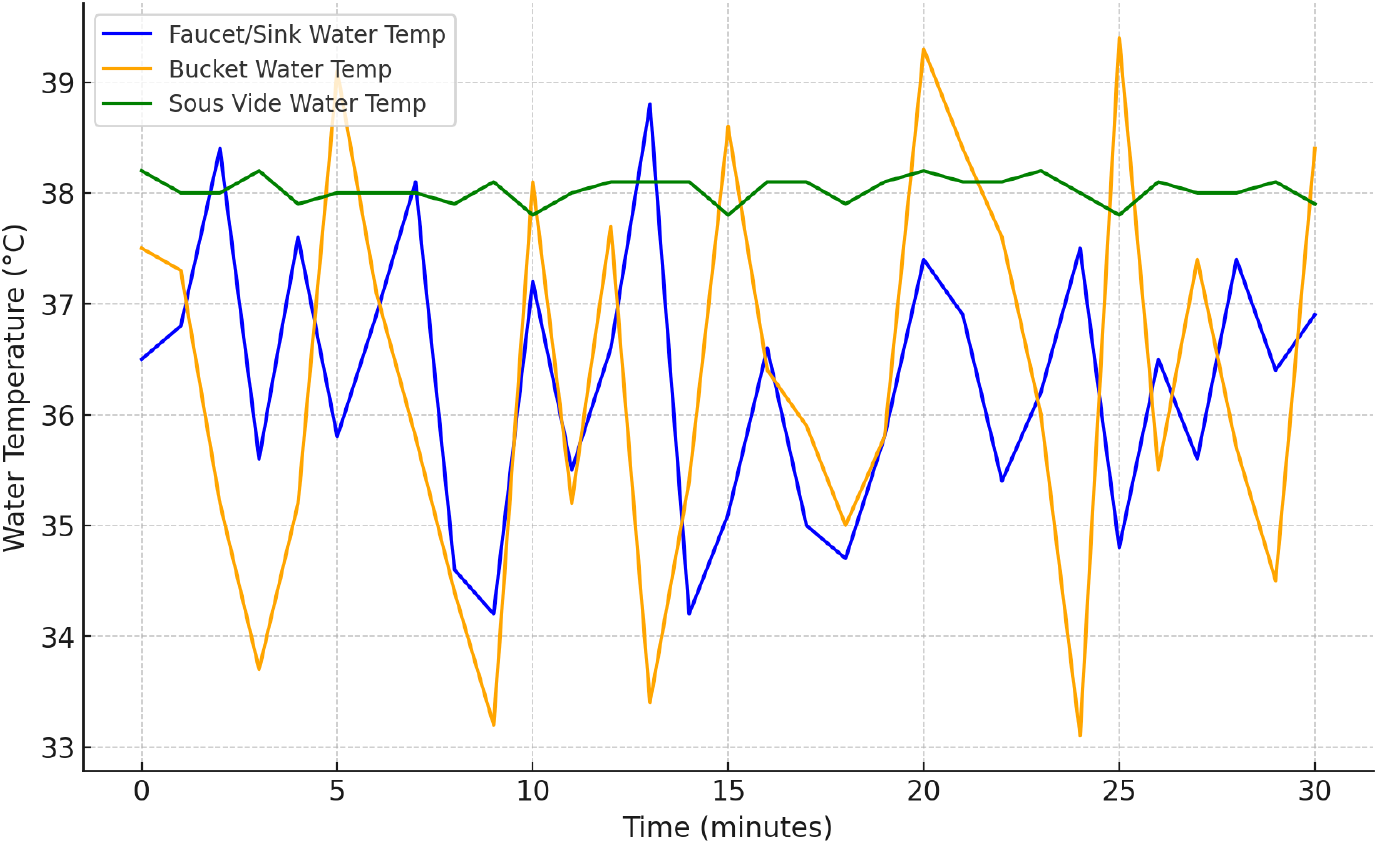
Water temperature variability over time for faucet/sink, bucket, and immersion circulator (sous vide) methods. *Note*. Mean water temperature data of 2 trials for each rewarming method.

### Summary of Key Findings

The immersion circulator method significantly outperformed the faucet/sink and bucket methods in terms of rewarming speed and consistency. It reached the target temperature in 9 minutes, compared to 22 minutes for the bucket and 29 minutes for the faucet/sink. Additionally, the immersion circulator method exhibited the least variability, maintaining a near-constant water temperature, while both the faucet/sink and bucket methods showed considerable fluctuations.

Lab personnel found it difficult to maintain bucket water in the target rewarming range. Additional hot water has to be obtained repeatedly. The bucket rewarming method had the highest workload, requiring continuous draining and refilling in order to maintain the target temperature. These results suggest that the immersion circulator method is the most effective and reliable technique for rewarming tissue in controlled laboratory conditions.

The statistical analysis of the three rewarming methods revealed differences in variability and overall rewarming efficiency. The immersion circulator method showed the least variability, with a coefficient of variation (CV) of just 0.31%, indicating highly stable temperature control. In comparison, the bucket method had a CV of 5.07%, and the faucet/sink method had a CV of 3.35%, both of which demonstrated considerably higher variability.

In terms of cumulative rewarming efficiency, as measured by the area under the curve (AUC), the immersion circulator outperformed the other methods, achieving an AUC of 1140.85. This was higher than the faucet/sink method’s AUC of 1088.3 and the bucket method’s AUC of 1087.35, indicating that the immersion circulator delivered heat more efficiently over the rewarming period.

These results demonstrate that the immersion circulator method is not only the most consistent in maintaining stable temperatures but also the most efficient in heat delivery.

## Discussion

### Interpretation

The results of this study clearly demonstrate that the immersion circulator method is the most effective technique for rewarming frostbitten tissue. The immersion circulator system achieved the target temperature of 38°C in the shortest time (∼9 minutes) and maintained near-perfect consistency, with no temperature fluctuations below 37°C and no additional workload on lab personnel. In contrast, both the faucet/sink and bucket methods showed significant variability, with water temperatures falling below 37°C for 70% and 76.7% of the rewarming period, respectively. For the bucket method, maintaining a stable rewarming temperature was extremely difficult. Moreover, for the sink method, the tap water temperature was highly variable over the course of the trial.

These findings suggest that immersion circulators offer superior temperature control, which is critical for preventing further tissue damage during rewarming. The key takeaway is that precise and consistent temperature management, as achieved with the immersion circulator method, is paramount in frostbite care.

### Previous Studies

Research on frostbite rewarming methods has largely focused on traditional techniques such as warm water immersion in sinks or buckets, as well as dry rewarming techniques like radiant heat or heat packs. Studies and reviews by Imray et al. (2009) and McIntosh et al. (2011) demonstrated that rapid rewarming in water between 37°C and 39°C is critical for reducing tissue damage. However, maintaining temperature manually, as methods like faucet or bucket immersion often lead to unstable temperatures, increasing the risk of refreezing or thermal injury (Handford et al., 2014).

The contribution of this study lies in its investigation of immersion circulator technology for frostbite treatment, an area that has been underexplored in the literature. Immersion circulator devices provide highly accurate temperature control, which traditional methods lack. By minimizing temperature fluctuations during rewarming, immersion circulators may help mitigate the risks of refreezing and thermal injury, improving overall tissue outcomes.

Furthermore, they do not require continuous effort by the healthcare worker to add tap water or drain and refill a bucket or basin. Our findings are consistent with a published case report by Daniel et al. (2022) where an immersion circulator was successfully used with minimal workload or variation in rewarming water temperature.

### Strengths and Limitations

One of the strengths of this study is the use of objective, continuous temperature measurements from both the water and tissue, which allowed us to accurately track the rewarming process and identify variability between methods. Additionally, the use of a controlled laboratory setting ensured consistency across trials, reducing the potential for confounding variables. However, the study has some limitations. First, the use of porcine legs, while appropriate for simulating human tissue rewarming, does not fully replicate the complexity of frostbite treatment in live patients, where vascular responses and immune reactions play crucial roles. Additionally, the study was conducted under ideal, controlled conditions, which may not fully represent real-world variability in clinical or field settings. The manual nature of the faucet and bucket methods introduces additional variables such as water supply temperature variability and human error, which may not be fully accounted for in the experimental setup.

### Clinical Implications

The clinical implications of this study are significant for both hospital and field management of frostbite. Rapid, controlled rewarming is essential to minimize tissue loss and improve long-term function, and immersion circulator technology offers several advantages over traditional rewarming methods in achieving this. In clinical settings, particularly in emergency departments, immersion circulator devices could provide a reliable and efficient method for maintaining precise rewarming temperatures, reducing the need for constant monitoring and manual adjustments, which are required with traditional methods (Handford et al., 2014).

In field settings, such as wilderness expeditions or military operations, where frostbite risk is high, portable immersion circulator devices could offer a valuable tool for first responders. By providing stable, controlled rewarming, they could reduce the risk of refreezing, thermal injury, and subsequent complications, which are common in manual methods. Field-deployable versions of immersion circulator devices could significantly enhance the care of frostbite patients in remote or resource-limited environments.

### Research Implications

This study highlights several important avenues for future research. First, clinical trials are needed to assess the efficacy of immersion circulator technology in real-world frostbite cases, particularly in terms of tissue preservation and functional recovery (Imray et al., 2009). While this study provides preliminary data from simulated rewarming scenarios, larger-scale clinical studies are necessary to confirm these findings and provide a basis for widespread adoption of immersion circulator technology in frostbite care.

Additionally, further research is needed to explore the balance between rapid rewarming and the risk of thermal injury or reperfusion damage. Studies like those by Cauchy et al. (2001), Poole and Gauthier (2016), and Poole et al. (2021) have shown that rapid rewarming is critical in the context of a complete care pathway including vasoactive vascular therapies, but there is limited research on the optimal rate of rewarming for different severities of frostbite.

Understanding the physiological effects of different rewarming speeds could lead to more refined treatment protocols, reducing the risk of complications while maximizing tissue preservation.

## Conclusion

In conclusion, this study demonstrates that the immersion circulator method offers a superior solution for the rewarming of frostbitten tissue, achieving faster and more consistent rewarming than traditional faucet or bucket methods. By providing precise temperature control, the immersion circulator method mitigates the risks associated with temperature variability, such as refreezing or overheating, and could lead to improved patient outcomes in frostbite care.

These findings have important implications for both clinical practice and future research, highlighting the potential for immersion circulator technology to enhance frostbite treatment protocols, particularly in settings where consistent rewarming is difficult to achieve.

## Data Availability

All data produced in the present work are contained in the manuscript.

## Acknowledgements

Tony Campanio and John Desouza, laboratory research assistants.

## Raw Data

| Time (min) | 1 Sink Water Temp (°C) | 1a Pig Leg Temp (°C) | 1b Pig Leg Temp (°C) | 2 Bucket Water Temp (°C) | 2a Pig Leg Temp (°C) | 2b Pig Leg Temp (°C) | 3 Immersion Circulator Water Temp (°C) | 3a Pig Leg Temp (°C) | 3b Pig Leg Temp (°C) |
| --- | --- | --- | --- | --- | --- | --- | --- | --- | --- |
| 0 | 36.5 | -7 | -6.8 | 37.5 | -7 | -6.7 | 38.2 | -7 | -7.1 |
| 1 | 36.8 | -6.2 | -5.9 | 37.3 | -5.2 | -5.7 | 38 | -1.4 | -1.4 |
| 2 | 38.4 | -5.4 | -5 | 35.2 | -3.5 | -3.6 | 38 | 4.2 | 4.4 |
| 3 | 35.6 | -4.7 | -4.9 | 33.7 | -1.8 | -2.1 | 38.2 | 9.9 | 10 |
| 4 | 37.6 | -3.9 | -3.6 | 35.2 | 0 | 0.2 | 37.9 | 15.5 | 15.5 |
| 5 | 35.8 | -3.1 | -2.6 | 39.1 | 0 | 0.4 | 38 | 21.1 | 21 |
| 6 | 36.9 | -2.3 | -2.6 | 37.1 | 1.5 | 1.8 | 38 | 26.8 | 26.7 |
| 7 | 38.1 | -1.6 | -1.2 | 35.8 | 3 | 3.3 | 38 | 32.4 | 32.4 |
| 8 | 34.6 | -0.8 | -0.6 | 34.4 | 4.6 | 4.3 | 37.9 | 38 | 37.8 |
| 9 | 34.2 | 0 | -0.5 | 33.2 | 6.1 | 6.2 | 38.1 | 38 | 38 |
| 10 | 37.2 | 0 | -0.1 | 38.1 | 7.6 | 8.1 | 37.8 | 38 | 37.9 |
| 11 | 35.5 | 1.9 | 1.9 | 35.2 | 9.1 | 8.9 | 38 | 38 | 37.9 |
| 12 | 36.6 | 3.8 | 3.7 | 37.7 | 10.6 | 11.1 | 38.1 | 38 | 38 |
| 13 | 38.8 | 5.7 | 5.2 | 33.4 | 12.2 | 12 | 38.1 | 38 | 37.9 |
| 14 | 34.2 | 7.6 | 7.4 | 35.4 | 13.7 | 14.1 | 38.1 | 38 | 38 |
| 15 | 35.1 | 9.5 | 9.3 | 38.6 | 15.2 | 14.7 | 37.8 | 38 | 38.2 |
| 16 | 36.6 | 11.4 | 11.2 | 36.4 | 16.7 | 16.7 | 38.1 | 38 | 37.8 |
| 17 | 35 | 13.3 | 13 | 35.9 | 18.2 | 18.4 | 38.1 | 38 | 38.1 |
| 18 | 34.7 | 15.2 | 15.5 | 35 | 19.8 | 20.2 | 37.9 | 38 | 37.9 |
| 19 | 35.8 | 17.1 | 17 | 35.8 | 21.3 | 20.8 | 38.1 | 38 | 37.8 |
| 20 | 37.4 | 19 | 18.6 | 39.3 | 22.8 | 23.2 | 38.2 | 38 | 37.8 |
| 21 | 36.9 | 20.9 | 20.7 | 38.4 | 24.3 | 24.3 | 38.1 | 38 | 37.9 |
| 22 | 35.4 | 22.8 | 23.3 | 37.6 | 25.8 | 26.2 | 38.1 | 38 | 38.1 |
| 23 | 36.2 | 24.7 | 24.9 | 36 | 27.4 | 27.4 | 38.2 | 38 | 37.9 |
| 24 | 37.5 | 26.6 | 26.7 | 33.1 | 28.9 | 29.3 | 38 | 38 | 38 |
| 25 | 34.8 | 28.5 | 28.3 | 39.4 | 30.4 | 30.1 | 37.8 | 38 | 38.2 |
| 26 | 36.5 | 30.4 | 30 | 35.5 | 31.9 | 31.4 | 38.1 | 38 | 37.8 |
| 27 | 35.6 | 32.3 | 32.6 | 37.4 | 33.4 | 33.1 | 38 | 38 | 37.8 |
| 28 | 37.4 | 34.2 | 34.3 | 35.7 | 35 | 34.9 | 38 | 38 | 38 |
| 29 | 36.4 | 36.1 | 35.7 | 34.5 | 36.5 | 36.9 | 38.1 | 38 | 38.1 |
| 30 | 36.9 | 38 | 38.3 | 38.4 | 38 | 38.3 | 37.9 | 38 | 37.9 |

